# Life Expectancy Loss among Native Americans During the COVID-19 Pandemic

**DOI:** 10.1101/2022.03.15.22272448

**Authors:** Noreen Goldman, Theresa Andrasfay

## Abstract

**Background:** There has been little systematic research on the mortality impact of COVID-19 in the Native American population.

**Objective:** We provide estimates of loss of life expectancy in 2020 and 2021 for the Native American population.

**Methods:** We use data on age-specific all-cause mortality rates from CDC WONDER and the 2019 life table recently released by the National Vital Statistics System for Native Americans to calculate life tables for the Native American population in 2020 and 2021 and obtain estimates of life expectancy reductions during the COVID-19 pandemic.

**Results:** The pandemic has set Native Americans further behind other major racial/ethnic groups in terms of life expectancy: the estimated loss in life expectancy at birth for Native Americans is 4.5 years in 2020 and 6.4 years in 2021.

**Conclusions:** These results underscore the disproportionate share of deaths experienced by Native Americans: a loss in life expectancy at birth in 2020 that is more than three years larger than that for Whites and about 1.5 years greater than the losses for the Black and Latino populations. Despite a successful vaccination campaign among Native Americans, the estimated loss in life expectancy at birth in 2021 unexpectedly exceeds that in 2020.

**Contribution:** The increased loss in life expectancy in 2021, despite higher vaccination rates than in other racial/ethnic groups, highlights the huge challenges faced by Native Americans in their efforts to control the deleterious consequences of the pandemic.

## Introduction

Despite heightened media attention to the large number of Native Americans dying from COVID-19, there has been little systematic research on the mortality impact of the coronavirus in this population. One exception is a recent paper demonstrating that age-standardized COVID-19 mortality rates among Native Americans exceeded the corresponding rates for the White, Black and Latino populations during 2020 (Leggat-Barr, Uchikoshi, and Goldman 2021). A separate study based on similar data, in this case for six racial and ethnic groups, indicated that the age-standardized COVID-19 death rate in 2020 among Native Americans was surpassed only by that for Hawaiian and other Pacific Islanders (Feldman and Bassett 2021).

In contrast to the standardized death rate used in these previous papers, measures of reductions in period life expectancy due to COVID-19 quantify the impact of the disease in a way that is easy to interpret while remaining independent of a population’s age distribution. Life expectancy at birth denotes the number of years newborns could expect to live if they experienced the age-specific death rates of a given period throughout their lives; similarly, life expectancy at age 65 reflects the expected number of years of life remaining for a person aged 65, again based on death rates for that period. The impact of COVID-19 on mortality of a population, be it a nation or a subgroup, can meaningfully be assessed in terms of declines in life expectancy between a period prior to and one during the pandemic.

Although the impact of COVID-19 on survival for the White, Black and Latino populations has been assessed in terms of declines in life expectancy (Andrasfay and Goldman 2021a, 2021b; Arias, Betzaida, et al. 2021; Woolf, Masters, and Aron 2021), this useful measure of longevity has not yet been estimated for Native Americans. The major obstacle has been the paucity of high-quality data. Measures of life expectancy are derived from life tables, which, until those published by the National Vital Statistics System (NVSS) in November 2021, had not been available for the total Native American population (Arias, Xu, et al. 2021). However, based on these recent pre-pandemic life tables and published classification ratios to correct for misreporting of AI/AN race, we are now able to calculate life tables for the pandemic period.

In this paper, we provide estimates of the loss of life expectancy in 2020 and 2021 for Native American men and women. Native Americans, who constitute about two percent of the US population, are defined as those self-identifying in the US Census as American Indian or Alaskan Native (AI/AN), excluding those also identifying as Latino. The analysis by Leggat-Barr and colleagues (2021) suggests that the mortality impact in 2020 was exceptionally large due to the many risk factors for viral exposure and viral severity present in this population: high poverty rates, crowded living arrangements, low access to quality healthcare driven in part by low rates of health insurance (aside from that provided by the poorly funded Indian Health Service (IHS)), frequent employment in frontline jobs, and a high prevalence of comorbidities that increase the risk of COVID-19 fatality. The estimated age-standardized COVID-19 death rate for AI/AN in 2020 exceeded those for other racial/ethnic groups nationally and in most of the states analyzed in their study. In addition, despite the implementation of mitigation strategies by Native American communities, such as contact tracing, sealing borders, mask mandates and enforcement of lockdowns (Foxworth et al. 2021), the state-level COVID-19 death rates in 2020 were strongly correlated with the proportion of Native Americans residing on reservations within the state (Leggat-Barr, Uchikoshi, and Goldman 2021).

To the best of our knowledge, no estimates of the overall mortality impact for Native Americans have been published for 2021. Despite the high burden of COVID-19 on the AI/AN population in 2020, there are reasons for optimism for 2021. From the start of vaccine availability, the AI/AN population has had higher rates of vaccine uptake than other racial and ethnic groups (Foxworth et al. 2021). Vaccination efforts that were culturally sensitive, combined with a steady supply of vaccine doses, were instrumental to this achievement. Most notable among these strategies was the use of vaccinated community elders as role models who emphasized the importance of preserving AI/AN culture and protecting Native American communities (Foxworth et al. 2021; Silberner 2021).

To assess the impact of COVID-19 on life expectancy in 2020 and 2021, we use the newly published 2019 life tables for the Native American population to provide information on death rates and life expectancy just prior to the pandemic (Arias, Xu, et al. 2021). These life tables have been adjusted by NVSS via linkages between the 2010 Census and death registration data to correct for misclassification of race and ethnicity on death certificates, a well-recognized and serious source of bias in data for the Native American population that has resulted in underestimation of mortality (Arias, Xu, et al. 2021; Espey et al. 2014; Jim et al. 2014). Incorporating these NVSS adjustments, we use age-specific all-cause mortality rates from 2020 and 2021 for Native Americans to estimate the loss in life expectancy at birth and at age 65 between 2019 and 2020 and between 2019 and 2021 for the Native American population as a whole and separately by sex. These estimates are then compared with published provisional life expectancy estimates for the White, Black, and Latino populations in 2020.

## Data and Methods

Counts of deaths from all causes and estimated population sizes by age, sex, race, and ethnicity in 2020 and 2021 were obtained from the CDC WONDER (Wide-ranging Online Data for Epidemiologic Research) database as of May 25, 2022 (CDC WONDER 2022). Deaths for 2020 are considered final while deaths for 2021 are considered provisional and therefore subject to reporting and processing delays.

We first adjust the all-cause counts of deaths in 2020 and 2021 to account for underreporting of Native American race on death certificates. We apply the age-group and sex-specific classification ratios published by NVSS (Arias, Xu, et al. 2021) to these death counts, with the assumption that the degree and age pattern of misclassification in 2020 and 2021 is equivalent to that used by NVSS in 2019. The average NVSS classification ratio was 1.34, with slightly higher values for males than females and the largest values at ages 45-74. We also borrow from the NVSS 2019 life tables the 1a0 values, which are the average numbers of person-years lived by infants who died before their first birthday. We use the adjusted death counts to estimate age-specific mortality rates (_n_M_x_) for both years and, subsequently, estimate the remaining life table columns using standard life table relationships (Preston, Heuveline, and Guillot 2000). For comparison, we present published life expectancy values for 2019 (Arias and Xu 2022) and provisional estimates for 2020 (Arias, Betzaida, et al. 2021) for the total US population and the Latino, White, and Black populations.

## Results

Simple descriptive statistics of COVID-19 deaths among Native Americans provide insights into changes in life expectancy during the pandemic. Before adjustment for racial misclassification, there were 4,265 deaths to Native Americans in 2020 and 4,615 in 2021 for which COVID-19 was the underlying cause of death. The median ages of these deaths declined from 69 to 65 years over this period, which, combined with the increase in numbers of deaths, suggests, contrary to expectation, that the loss in life expectancy at birth may be greater in 2021.

Brought about partly by the younger age distribution of the Native American than the White population (Akee and Reber 2021; Leggat-Barr, Uchikoshi, and Goldman 2021), the median ages of COVID-19 deaths are more than a decade higher for Whites – 82 in 2020 and 75 in 2021 – than for Native Americans. However, as shown in Figure 1, these differences in ages at death primarily reflect the much higher COVID-19 mortality rates among Native Americans at younger ages. COVID-19 death rates in the young adult and middle age range are about ten or more times as high for Native Americans as for Whites in 2020, and are reduced to about four to five times as high in 2021. Above age 75, death rates are approximately twice as high among Native Americans in both years. Preliminary work suggests that the Black and Latino populations also experienced reductions in age-specific death rates relative to Whites from 2020 to 2021 driven by rising mortality rates among Whites (Andrasfay and Goldman 2022).

**Figure 1:**
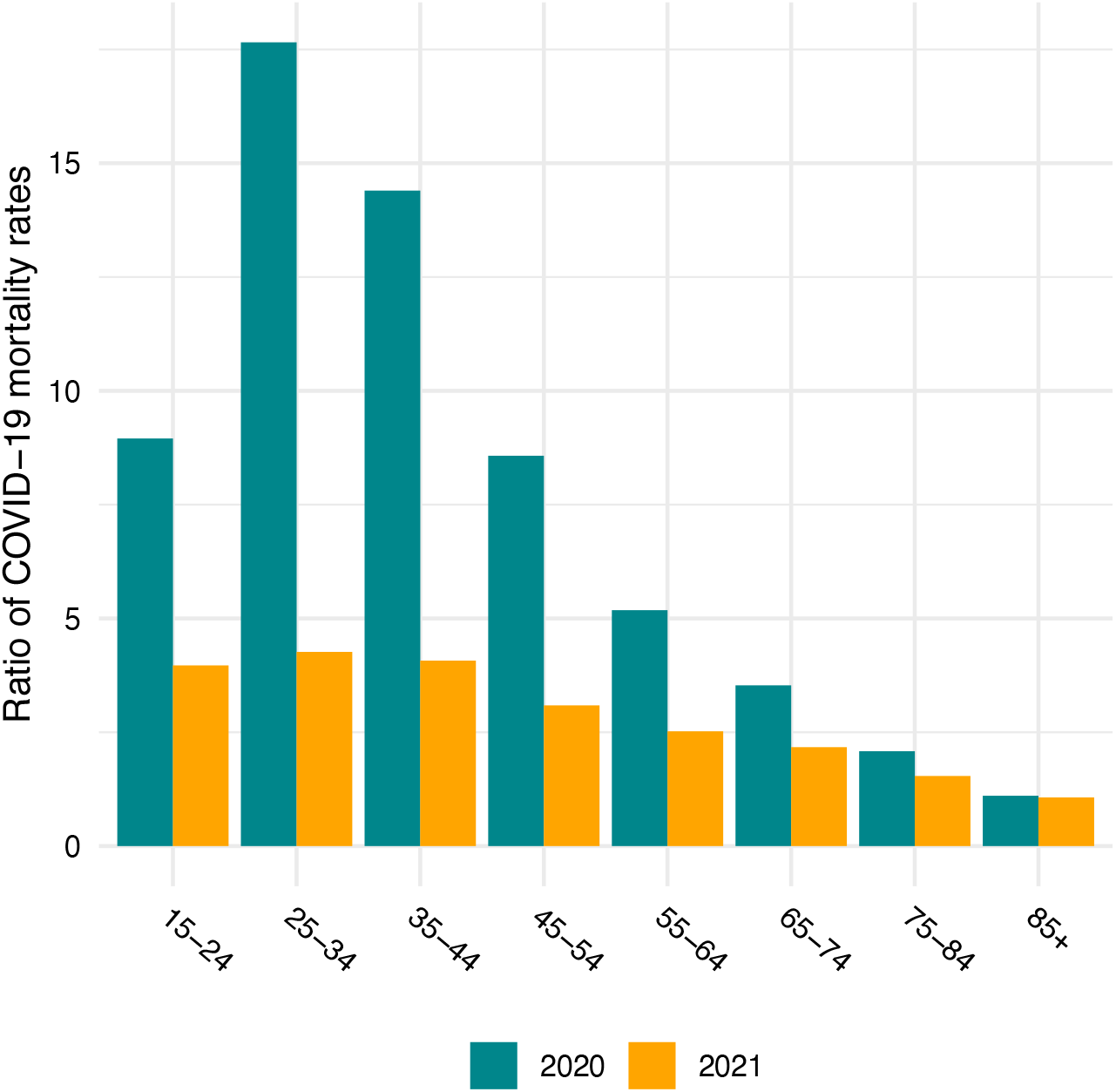
Ratio of Native American to White COVID-19 age-specific mortality rates in 2020 and 2021. Data are from CDC WONDER and include all deaths for which COVID-19 is the underlying cause of death. Native American COVID-19 death counts have been adjusted to account for misreporting of AI/AN race on death certificates. Ratios below age 15 are not shown because counts of COVID-19 deaths are suppressed at these ages for confidentiality.

Table 1 presents life expectancy at birth (e_0_) and at age 65 (e_65_) by sex as calculated by NVSS in their 2019 life tables along with our results for 2020 and 2021. The estimated declines in life expectancy at birth are 4.5 years in 2020 and a substantially larger estimate of 6.4 years in 2021. The pandemic reduced Native American life expectancy at birth from the already low value of 72 years in 2019 to about 67 years in 2020 and about 65 years in 2021. The larger reductions for men than women have resulted in a gender gap in life expectancy at birth estimated at 7.2 years in 2021 compared with 6.4 years in 2019 and are consistent with larger reductions seen among men in other populations in the US. The estimated impact of the pandemic on e_65_ is about two years in each of 2020 and 2021, slightly higher for women than men in contrast to the gender difference in loss of life expectancy at birth.

**Table 1:**
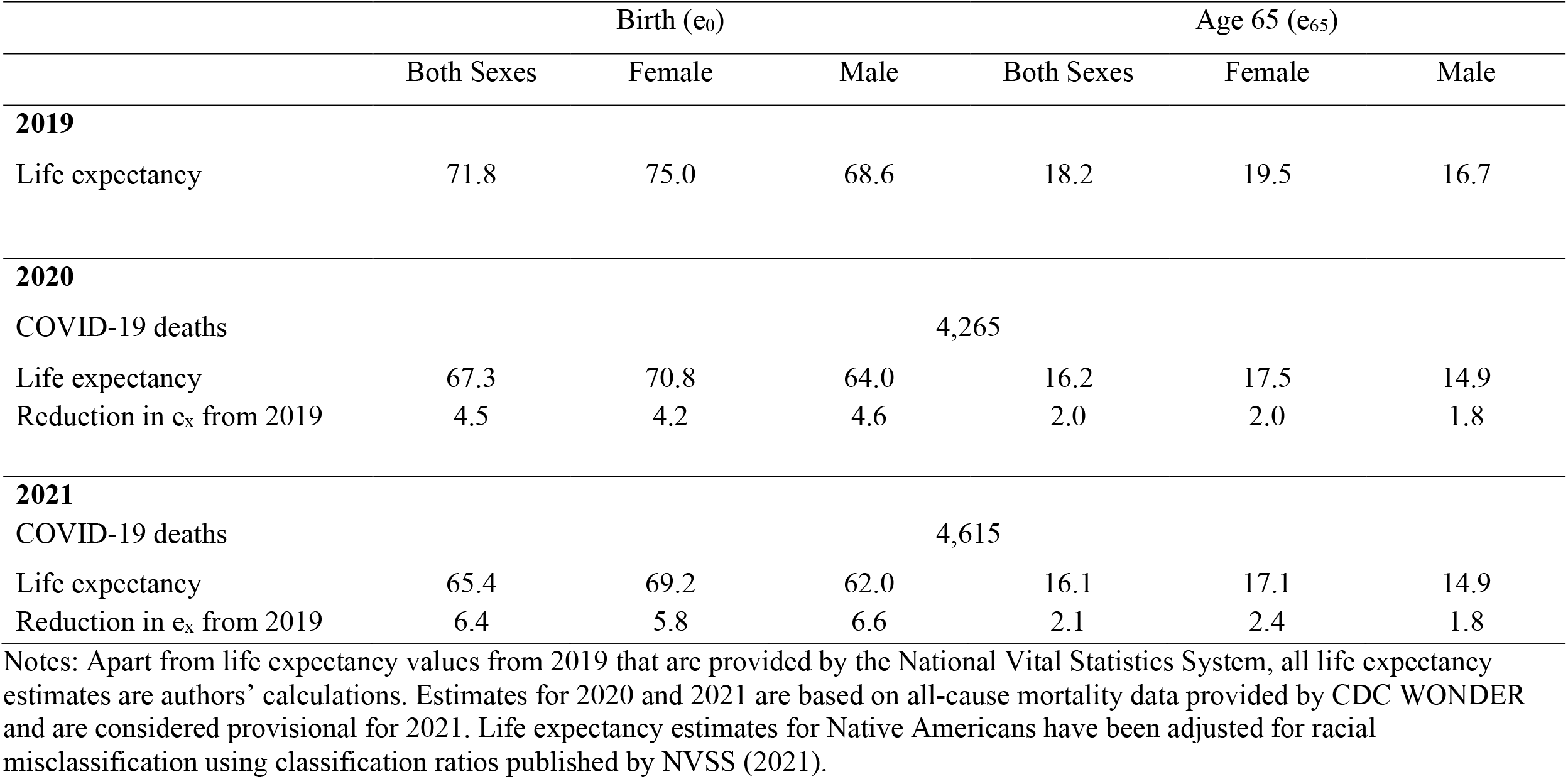
Estimates of life expectancy and losses in life expectancy for the Native American population.

Figure 2 compares the estimated loss in life expectancy by sex between 2019 and 2020 across racial and ethnic groups, based on our estimates for Native Americans and those from Arias et al. (2021) for the Latino, White, and Black populations. This figure shows that, even before the pandemic, the Native American population had much lower life expectancy than other major racial/ethnic groups in the US. The pre-pandemic (2019) e_0_ of 71.8 for the AI/AN population was seven years below that of the White population and a decade lower than that of the Latino population. The pandemic has set the AI/AN population behind even further: the estimated loss in e_0_ for Native American men and women in 2020 greatly exceeds those for all the other groups. For both sexes combined, the loss in e_0_ for Native Americans of 4.5 years is over three years greater than that for Whites and about 1.5 years larger than those for the Black and Latino populations. The two-year loss in e_65_ for Native Americans (both sexes combined) also exceeds the corresponding declines for the other racial/ethnic groups. Only Latino men experienced a greater loss in e_65_ than Native American men.

**Figure 2:**
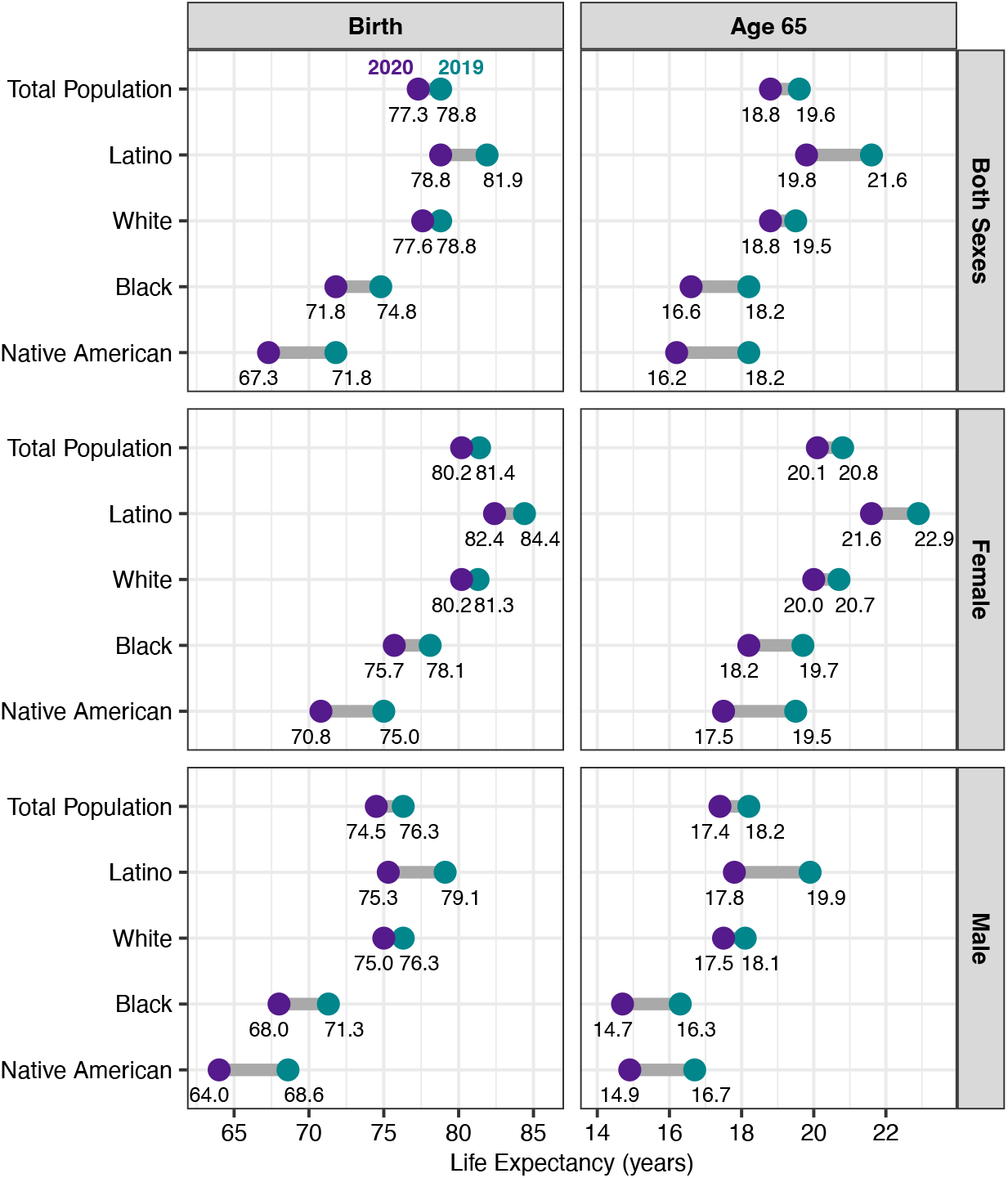
Life expectancy at birth and at age 65 by race/ethnicity, estimates for 2020 compared to 2019. Life expectancy values for the Total, Latino, White, and Black populations are taken from published NVSS provisional estimates. Estimated 2020 life expectancy values for the Native American population are authors’ calculations from CDC WONDER mortality data and incorporate adjustments for racial misclassification of deaths to Native Americans.

## Discussion

Throughout the pandemic, scholars and journalists have acknowledged the disproportionate share of COVID-19 deaths experienced by Native Americans. Here, we quantified this mortality burden in terms of reductions in life expectancy at birth and at age 65 in a way that takes into account both deaths from COVID-19 and excess deaths from other causes. We estimate that life expectancy at birth in 2020 and 2021 has declined to approximately 67 and 65 years respectively, shockingly low values in a high-income country. These levels of life expectancy are far below those in every country in the Americas with the sole exception of Haiti (where the estimated life expectancy is similar at 64), and several years below values observed prior to the pandemic in India, Pakistan and Nepal (Population Reference Bureau 2020). COVID-19 deaths have exacerbated an already stark mortality disparity between Whites and Native Americans in the US, augmenting a seven-year gap in life expectancy prior to the pandemic to 10 years in 2020. Our estimates suggest that reductions in 2020 life expectancy at birth were almost four times as large for Native Americans as they were for Whites. Although much of this decline resulted directly from COVID-19 deaths, mortality from non-COVID-19 causes also increased substantially between 2019 and 2021: death rates from all non-COVID-19 causes combined rose progressively through the three years for all adult age groups, with the largest proportional increases between ages 25 and 44 (authors’ calculations from CDC WONDER, 2022).

One limitation of this analysis is that our estimates for 2021 are based on provisional counts of death, as are the comparisons for other racial/ethnic groups in 2020. A second limitation is that our estimates of life expectancy for 2020 and 2021 depend heavily on the NVSS classification ratios that were derived from the matching of census and death certificate information a decade earlier (2010-2011). The adjustment factor (1.34 on average) results primarily from underreporting of Native American race on death certificates, with most misclassified Native American decedents reported as White on death certificates (Arias, Xu, et al. 2021). Such misclassification has been especially prevalent for persons self-identifying with multiple races in census data, particularly among AI/AN residing outside of Indian Health Service contract delivery areas (Jim et al. 2014). To the extent that misclassification of race/ethnicity has changed over the past decade, these estimates of life expectancy will be biased (e.g., too low if underreporting of Native American race on death certificates has diminished). However, because we use the same classification ratios in 2019, 2020, and 2021, our estimates of *loss* in life expectancy over this period are likely robust to the extent that these ratios have remained constant over the three years.

The increase in mortality rates during the pandemic among Native Americans was likely due in large part to the high rates of chronic disease in this population prior to the pandemic. The high prevalence of co-morbidities in the AI/AN population not only increased susceptibility to COVID-19 infection and severity, but COVID-19 infection and potential experiences with “long COVID” likely increased the severity and hence fatality from non-COVID-19 illnesses due to the physiological sequelae of the infection and reduced availability and use of healthcare during the pandemic (Leggat-Barr, Uchikoshi, and Goldman 2021). As with the US population at large, it is also likely that detrimental health-related behaviors, such as smoking, drinking and drug use, became more frequent during this time (Kalkhoran, Levy, and Rigotti 2021; Zhang et al. 2021). There is already evidence of a more than 40% increase in fatal drug overdoses among Native Americans in 2020, which exceeded the increases for Whites and Latinos (Friedman and Hansen 2022; Hedegaard et al. 2021).

Despite the successful vaccination campaign among Native Americans, as well as efforts to increase COVID-19 testing and modernize the health IT infrastructure of the IHS (Indian Health Service 2022), the estimated loss in life expectancy at birth in 2021 substantially exceeds that in 2020. There are several plausible explanations for this unexpected and disturbing finding. First, because the initial vaccine rollout prioritized healthcare workers, no racial/ethnic group had substantial vaccination coverage during January-February 2021 – two of the deadliest months of the pandemic (Centers for Disease Control and Prevention 2022; Ndugga et al. 2021). Second, two highly contagious variants that partially evaded vaccine-acquired and natural immunity, Delta and Omicron, emerged in 2021 (Kupferschmidt and Wadman 2021; del Rio, Omer, and Malani 2021). Third, the Native American population, like the general US population, still has a substantial proportion of unvaccinated persons, especially among younger adults, and a relatively low uptake of booster doses (Centers for Disease Control and Prevention 2022). Fourth, death rates from several chronic conditions and drug overdoses increased during the pandemic and these increases did not abate in 2021. Fifth, and most importantly, Native Americans continue to experience large social, economic and health inequities, some of which have persisted for centuries. These factors increase risks of COVID-19 infection, hospitalization and death: high rates of poverty, poor housing infrastructure including inadequate plumbing, crowded living conditions often involving multigenerational families, employment in low-income frontline jobs that cannot be performed remotely, high rates of co-morbidities (particularly obesity and diabetes) that increase the severity and fatality of COVID-19, a high prevalence of smoking, low rates of health insurance other than the IHS, and poorly resourced healthcare systems that provide inadequate and often inaccessible care (Leggat-Barr, Uchikoshi, and Goldman 2021). Because many of these risk factors are more prevalent in tribal lands than in non-tribal areas, vulnerability to COVID-19 is especially high for Native Americans residing on reservations (Leggat-Barr, Uchikoshi, and Goldman 2021).

Although COVID-19 death rates among Native Americans undoubtedly would have been larger in the absence of a successful vaccine campaign, our results underscore the huge challenges faced by this population in their efforts to control the appalling consequences of the ongoing pandemic that coexist with high rates of morbidity and mortality from other causes. The large financial investment in the American Rescue Plan to enhance identification and treatment of COVID-19 infections and strengthen the public health infrastructure for the Native American population is a significant step forward (Indian Health Service 2021).

## Data Availability

The data used in this study are all publicly available through the provided links.

https://wonder.cdc.gov/

## Acknowledgments

We would like to thank Katherine Leggat-Barr, Fumiya Uchikoshi, and the anonymous referees for helpful comments. This study was supported in part by the National Institute on Aging (grant no. T32AG000037).

## References

Akee, R. and Reber, S. (2021). American Indians and Alaska Natives Are Dying of COVID-19 at Shocking Rates. Washington, D.C.: The Brookings Institution. https://www.brookings.edu/research/american-indians-and-alaska-natives-are-dying-of-covid-19-at-shocking-rates/.

Andrasfay, T. and Goldman, N. (2021a). Association of the COVID-19 Pandemic With Estimated Life Expectancy by Race/Ethnicity in the United States, 2020. JAMA Network Open 4(6):e2114520–e2114520. doi:10.1001/jamanetworkopen.2021.14520.

Andrasfay, T. and Goldman, N. (2021b). Reductions in 2020 US life expectancy due to COVID-19 and the disproportionate impact on the Black and Latino populations. Proceedings of the National Academy of Sciences 118(5). doi:10.1073/pnas.2014746118.

Andrasfay, T. and Goldman, N. (2022). Reductions in US life expectancy from COVID-19 by Race and Ethnicity: Is 2021 a repetition of 2020? :2021.10.17.21265117. doi:10.1101/2021.10.17.21265117.

Arias, E., Betzaida, T.-V., Ahmad, F., and Kochanek, K. (2021). Provisional Life Expectancy Estimates for 2020. Hyattsville, MD: National Center for Health Statistics (U.S.). Vital Statistics Rapid Release. doi:10.15620/cdc:107201.

Arias, E. and Xu, J. (2022). United States Life Tables, 2019. National Vital Statistics Reports 70(19). https://www.cdc.gov/nchs/data/nvsr/nvsr70/nvsr70-19.pdf.

Arias, E., Xu, J., Curtin, S., Bastian, B., and Tejada-Vera, B. (2021). Mortality Profile of the Non-Hispanic American Indian or Alaska Native Population, 2019. National Vital Statistics Reports(12):1–27.

CDC WONDER (2022). Provisional Mortality on CDC WONDER Online Database. [electronic resource]. https://wonder.cdc.gov/.

Centers for Disease Control and Prevention (2022). COVID-19 Cases, Deaths, and Trends in the US | CDC COVID Data Tracker [electronic resource]. https://covid.cdc.gov/covid-data-tracker.

Espey, D.K., Jim, M.A., Richards, T.B., Begay, C., Haverkamp, D., and Roberts, D. (2014). Methods for Improving the Quality and Completeness of Mortality Data for American Indians and Alaska Natives. American Journal of Public Health 104(S3):S286–S294. doi:10.2105/AJPH.2013.301716.

Feldman, J.M. and Bassett, M.T. (2021). Variation in COVID-19 Mortality in the US by Race and Ethnicity and Educational Attainment. JAMA Network Open 4(11):e2135967. doi:10.1001/jamanetworkopen.2021.35967.

Foxworth, R., Redvers, N., Moreno, M.A., Lopez-Carmen, V.A., Sanchez, G.R., and Shultz, J.M. (2021). Covid-19 Vaccination in American Indians and Alaska Natives — Lessons from Effective Community Responses. New England Journal of Medicine 385(26):2403–2406. doi:10.1056/NEJMp2113296.

Friedman, J.R. and Hansen, H. (2022). Evaluation of Increases in Drug Overdose Mortality Rates in the US by Race and Ethnicity Before and During the COVID-19 Pandemic. JAMA Psychiatry. doi:10.1001/jamapsychiatry.2022.0004.

Hedegaard, H., Miniño, A.M., Spencer, M.R., and Warner, M. (2021). Drug Overdose Deaths in the United States, 1999-2020. Hyattsville, MD: National Center for Health Statistics (U.S.). NCHS data brief. https://www.cdc.gov/nchs/products/databriefs/db428.htm.

Indian Health Service (2021). Biden Administration Invests Additional $1.8 Billion in American Rescue Plan Funding to Combat COVID-19 in Indian Country. Indian Health Service. Indian Health Service Press Release. https://www.ihs.gov/sites/newsroom/themes/responsive2017/display_objects/documents/Press-Release-Additional-1.8-billion-in-ARP-funding-6162021.pdf.

Indian Health Service (2022). Coronavirus | Indian Health Service (IHS). Rockville, MD: Indian Health Service. https://www.ihs.gov/coronavirus/.

Jim, M.A., Arias, E., Seneca, D.S., Hoopes, M.J., Jim, C.C., Johnson, N.J., and Wiggins, C.L. (2014). Racial Misclassification of American Indians and Alaska Natives by Indian Health Service Contract Health Service Delivery Area. American Journal of Public Health 104(S3):S295–S302. doi:10.2105/AJPH.2014.301933.

Kalkhoran, S.M., Levy, D.E., and Rigotti, N.A. (2021). Smoking and E-Cigarette Use Among U.S. Adults During the COVID-19 Pandemic. American Journal of Preventive Medicine. doi:10.1016/j.amepre.2021.08.018.

Kupferschmidt, K. and Wadman, M. (2021). Delta variant triggers new phase in the pandemic. Science 372(6549):1375–1376. doi:10.1126/science.372.6549.1375.

Leggat-Barr, K., Uchikoshi, F., and Goldman, N. (2021). COVID-19 risk factors and mortality among Native Americans. Demographic Research 45:1185–1218.

Ndugga, N., Pham, O., Hill, L., Artiga, S., and Parker, N. (2021). Latest Data on COVID-19 Vaccinations by Race/Ethnicity. San Francisco, CA: Kaiser Family Foundation. https://www.kff.org/coronavirus-covid-19/issue-brief/latest-data-on-covid-19-vaccinations-race-ethnicity/.

Population Reference Bureau (2020). 2020 World Population Data Sheet. Washington, D.C.: Population Reference Bureau. https://www.prb.org/wp-content/uploads/2020/07/letter-booklet-2020-world-population.pdf.

Preston, S., Heuveline, P., and Guillot, M. (2000). Demography: Measuring and Modeling Population Processes. 2001.

del Rio, C., Omer, S.B., and Malani, P.N. (2021). Winter of Omicron—The Evolving COVID-19 Pandemic. JAMA. doi:10.1001/jama.2021.24315.

Silberner, J. (2021). Covid-19: How Native Americans led the way in the US vaccination effort. BMJ 374:n2168. doi:10.1136/bmj.n2168.

Woolf, S.H., Masters, R.K., and Aron, L.Y. (2021). Effect of the covid-19 pandemic in 2020 on life expectancy across populations in the USA and other high income countries: simulations of provisional mortality data. BMJ 373:n1343. doi:10.1136/bmj.n1343.

Zhang, X., Oluyomi, A., Woodard, L., Raza, S.A., Adel Fahmideh, M., El-Mubasher, O., Byun, J., Han, Y., Amos, C.I., and Badr, H. (2021). Individual-Level Determinants of Lifestyle Behavioral Changes during COVID-19 Lockdown in the United States: Results of an Online Survey. International Journal of Environmental Research and Public Health 18(8):4364. doi:10.3390/ijerph18084364.

